# Modelling the impact of initiation delay, duration and prior PrEP usage on the prophylactic efficacy of FTC/TDF-containing post-exposure prophylaxis

**DOI:** 10.1101/2024.07.22.24310798

**Authors:** Lanxin Zhang, Simon Collins, Julie Fox, Max von Kleist

**Author notes:** Corresponding author: Max von Kleist, Nordufer 20, Berlin, 13353, Germany, MvK. E-mail addresses of authors: LZ, SC, JF.

## Abstract

**Introduction:** Pre- and post-exposure prophylaxis (PrEP and PEP) are important pillars of the HIV prevention portfolio to reduce the risk of infection just before or after HIV exposure. While PrEP efficacy has been elucidated in many randomized clinical trials, corresponding data for PEP is extremely difficult to obtain in a controlled setting. Consequently, it is almost impossible to study the impact of PEP initiation delay and duration on HIV risk reduction clinically, which would inform recommendations on PEP use.

**Methods:** We employ pharmacokinetics, pharmacodynamics, and viral dynamics models, along with individual factors, such as drug adherence to investigate the impact of initiation delay and PEP duration on HIV risk reduction. We evaluated PEP using two- and three-drug regimens with a FTC/TDF backbone. Moreover, we study PEP efficacy in the context of PrEP-to-PEP transitions.

**Results:** In our simulations, early initiation of PEP emerged as a pivotal factor for HIV risk reduction. We found that 2-drug (FTC/TDF) PEP may insufficiently protect when initiated *>* 1 hour post-exposure. When adding a third drug, early initiation was still a critical factor, however, over 90% efficacy could be achieved when PEP was initiated 48hours post-exposure and taken for at least 14-28days, depending on the efficacy of the third-drug component. When investigating PrEP- PEP transitions, we observed that preceding PrEP can (i) contribute directly to prophylactic efficacy, and (ii) boost subsequent PEP efficacy by delaying initial viral dynamics and building-up drug concentrations, overall facilitating self-managed transitioning between PrEP and PEP.

**Conclusions:** Our study confirms the critical role of early (*<* 48hours) PEP initiation, preferably with three drugs taken for 28days. Self-start with TDF/FTC and later addition of a third drug is better than not self-starting. Furthermore, our study highlights the synergy between recent PrEP intake and PEP and may help to inform recommendations on PEP use.

## Introduction

The human immunodeficiency virus (HIV) remains a public health challenge with an estimated 1.3 million new infections in 2022 [1]. To date, with a handful of exceptions, HIV infection cannot be cured [2]. However, major successes in antiviral drug development allow to not only to prevent AIDS, but to suppress the virus to an extent where the treated individual is non-infectious [3, 4]. However, an HIV diagnosis needs to made and subsequent treatment currently needs to be taken life-long, which, in addition to individual burden, relies on HIV testing and treatment availability, medical care infrastructure and funding. HIV prevention through vaccination would constitute an ideal means to fight the pandemic. However, developing an effective HIV vaccine turned out to be extremely challenging, with all recent vaccine trials prematurely terminated due to failure in demonstrating clinical efficacy [5]. In the absence of effective vaccines, pre-exposure prophylaxis (PrEP) has partly taken its place. Four effective regimen are currently available: once daily emtricitabine (FTC) with either tenofovir disoproxil fumerate (TDF) or tenofovir alfenamide (TAF) can be administered orally, long-acting cabotegravir (CAB) can be injected every two month. Monthly dapivirine (DPV) vaginal rings to prevent infection through receptive vaginal intercourse recently received positive review by the European Medicines Agency (EMA). Twice-yearly injectable lenecapavir demonstrated potential in clinical phase III. Of the available PrEP options, oral TDF/FTC is widely available as a generic and rolled out in both low- and high-income countries.

Post-exposure prophylaxis (PEP) taken *after* suspected sexual-, or occupational exposure to HIV [6] denotes another important preventive measure to reduce infection risk. Current guidelines recommend to initiate oral PEP within 72hours after suspected virus exposure and to continue the regimen for 28 days [6–8]. National [6, 8] and international guidelines [9] differ with regards to recommending two- or three-drug regimens for PEP: For example, TDF/FTC + raltegravir or dolutegravir are recommended in the US, whereas the WHO 2014 guidelines also discuss scenarios where two-drug regimens with generics may be recommended. To date, TDF/FTC denotes the preferred backbone in PEP, whereas different choices of third-component drugs may be used [10]. However, because of operational and ethical challenges no randomized controlled trial has been conducted to test PEP efficacy directly. Current evidence for non-occupational PEP efficacy has been synthesized from animal transmission models, observational studies of health care workers receiving prophylaxis after occupational exposures, and observational- and case studies of PEP use [6, 8]. However, results from observational studies may be impacted by many factors such as individual adherence- and risk behavior [11] and differences in regards to the utilized PEP drugs [8]. Although the developed guidelines are based on impressive trans-disciplinary synthesis of evidence across heterogeneous data sources, it has not been possible to date to elucidate the sensitivity of particular PEP regimen to delays in initiation, PEP duration, as well as the impact of PrEP on PEP efficacy.

In the absence of randomized controlled trial data on PEP efficacy, mathematical modelling may support the synthesis of evidence, by integrating available knowledge on drug pharmacokinetics, as well as early viral dynamics. However, to our knowledge, no such modelling exists to date. By considering population pharmacokinetics, we extended a recently developed mathematical model [12] to analyse PEP efficacy for two- and three-drug regimens, and to test the impact of delays in ‘time to PEP’, as well as PEP duration. Finally, we investigate the transition from PrEP to PEP, providing a comprehensive understanding of the continuum preventive portfolio.

## Methods

We combined population pharmacokinetic models of oral FTC, TDF, EFV and DTG [13–16] with viral dynamics models [17, 18] and a novel numerical scheme [19] to estimate the prophylactic efficacy of PEP with a TDF/FTC backbone for any dosing pattern of interest, as well as various PrEP-to-PEP transitions. The overarching goal was to understand sensitivity of PEP efficacy towards timing, delay and duration of PEP with- and without prior PrEP administration.

### Prophylactic efficacy

In clinical trials, *average* HIV risk reduction is quantified in terms of incidence reduction in an intervention vs. a control arm [20–23]. In a mathematical model of within-host viral replication, the same quantity may be derived directly by computing the reduction of infection probability *per viral exposure* due to a prophylactic regimen *𝒮*:

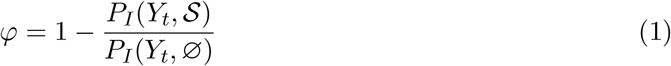

where *P*_*I*_(*Y*_*t*_, *𝒮*) and *P*_*I*_(*Y*_*t*_, ∅) denote the the infection probability in the presence- and absence of a prophylactic regimen 𝒮 upon exposure with *Y*_*t*_ viral particles at time *t*. Notably, the infection probability is the complement of the probability that the virus may eventually be eliminated in the exposed host, i.e. *P*_*I*_(*Y*_*t*_, *𝒮*) = 1 − *P*_*E*_(*Y*_*t*_, *𝒮*).

### Virus exposure model

We used previously developed exposure models for sex without condoms [24]. In these models, the number of infectious viruses (inoculum size *Y*_*t*_) that are transmitted to- and reaching an anatomical site where they may spark an infection, are estimated from a binomial distribution, *Y*_*t*_ ∼ *ℬ*(VL, *r*), where VL denotes the donor virus load, and the ‘success rate’ *r* depended on the type of exposure. Throughout this study, unless stated otherwise, we utilize the exposure model designed for receptive vaginal intercourse.

### HIV viral dynamics model

To compute the viral elimination probability in the exposed host for prophylactic regimen *𝒮*, we employ a within-host viral dynamics model [17, 18], depicted in Fig S1. The model considers replication of free infectious viruses, early- and productively infected T-cells, as well as long-lived cells such as macrophages and latently infected T-cells, which are believed to be an obstacle for the within-host clearance of HIV [25]. The model was derived from first principles [17] and allows to model pharmacodynamic effects of all antiviral classes [26]. Moreover, it allows to incorporate state-of-the-art population pharmacokinetic models.

### Pharmacokinetics

We used the previously developed pharmacokinetic models of emtricitabine (FTC) [13] and tenofovir disoproxil fumarate (TDF) [14], which allow to predict prodrug pharmacokinetics in blood plasma, as well as the pharmacokinetics of the active tri-phosphorylated moieties in peripheral blood mononuclear cells (PBMC). In line our recent findings [12], we assume that the concentration of tenofovir-diphosphate (TFV-DP) and emtricitabine-triphosphate (FTC-TP) in PBMCs predicts the prophylactic effect. We adopted recently developed PK models for dolutegravir (DTG) [15] and efavirenz (EFV) [27]. To capture the impact of individual pharmacokinetic variability, we sampled PK parameters for 1000 virtual patients per drug, utilizing distributions described in the aforementioned original sources. We considered once daily oral doses of 300/200mg, 50mg and 400mg for TDF/FTC, DTG and EFV.

### PK-PD link

The active intracellular components of TDF/FTC, i.e. TFV-DP and FTC-TP are nucleotides reverse transcriptase inhibitors (NRTI). To evaluate their combinatorial effect, we adopted a model for the molecular mechanism of action (MMOA) and drug-drug interaction [28]. For DTG and EFV, their direct effect can be modelled using the Emax equation [29], corrected by plasma protein binding, and was assumed to be additive with the TDF/FTC backbone.

### Numerics

We adopted the numerical scheme (PGS) from [19] to formulate a set of ordinary differential equations that allows computing extinction probabilities *P*_*E*_(*Y*_*t*_), *𝒮* of each compartment of the viral dynamics model, subject to pharmacokinetics and pharmacodynamics of the considered drugs, eq. (S16).

## Results

### ‘Time to PEP’ is the most critical parameter

Currently, the WHO recommends to initiate PEP up to 3 days after potential viral exposure and to continue PEP for 28 days [9]. Using our modelling framework, we evaluated how PEP initiation delay may alter prophylactic efficacy. As a first test case, we explored the efficacy of 2-drug (oral TDF/FTC) PEP, as these drugs may be available in many settings where PrEP is implemented. We created 1000 virtual individuals and simulated individual pharmacokinetics based on the dosing profiles in Fig 1A. Using the model, we then computed the prophylactic efficacy for each virtual individual, if a 2-drug PEP with daily TDF/FTC was initiated at different time points post viral exposure and taken for 28 days. Fig 1C (red line, grey areas) depicts summary statistics of derived PEP efficacy estimates across the cohort of virtual individuals (median, interquartile ranges and 95% confidence intervals). From the simulations, it is evident that ≥ 90% 2-drug PEP efficacy in only achieved if TDF/FTC is initiated within one hour after virus exposure. Efficacy steeply drops to *<* 50% when TDF/FTC-PEP was initiated 20hours after virus exposure. We also found that longer duration of 2-drug TDF/TFC PEP could not compensate for delayed initiation (Fig 1B, Fig 1D red line) with efficacy remaining low (median efficacy *<* 20%), when PEP was initiated 48hours after virus exposure and taken for up to 7 weeks. We tested whether a third drug component (DTG or EFV) may impact on prophylactic efficacy and change sensitivity to ‘time to PEP’ and ‘PEP duration’, Fig 1A–D. Compared to 2-drug PEP, 3-drug PEP provided *>* 88% protection against sexual transmission, when initiated 2days post-exposure and continued for 28days (Fig 1E). When initiated 2days post-exposure, we predicted that TDF/FTC + EFV provided *>* 90% HIV risk reduction when taken at least for 2 weeks, whereas TDF/FTC + DTG provided 85 − −90% HIV risk reduction when taken for at least 4 weeks, Fig 1E. In contrast to 2-drug PEP, we predicted that PEP efficacy with TDF/FTC + EFV or DTG increased with extended duration of PEP.

**Fig 1.**
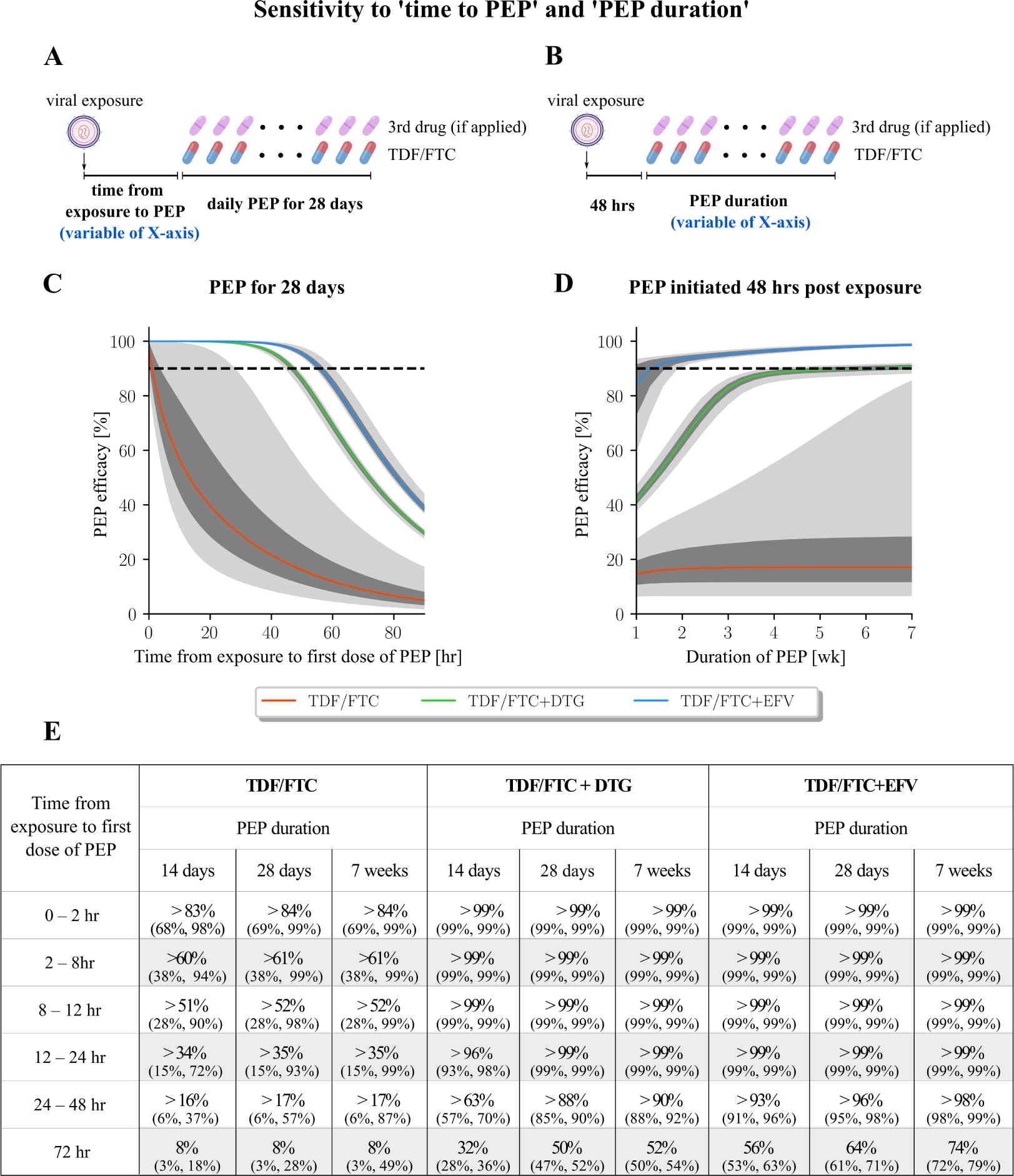
Sensitivity of TDF/FTC-based PEP on initiation delay and PEP duration. A & B: Schematic of the dosing regimen in panel C and D, respectively. C: PEP efficacy of TDF/FTC (red line), TDF/FTC + EFV (blue line), or TDF/FTC + DTG (green line) when initiated at different delays post virus exposure and taken for 28 days once-daily. D: Efficacy of TDF/FTC (red line), TDF/FTC + EFV (blue line) and TDF/FTC + DTG (green line) when initiated 48 hours post virus exposure and taken for different durations. E: Numerical results for different ‘times to PEP’, ‘PEP durations’ and regimen. Values denote the median efficacy and 95% confidence interval evaluated at the maximum ‘time to PEP’ of the indicated interval (e.g. 8hr for the 2-8hr interval). All computations were conducted on 1000 virtual patients. The daily oral dose for each drug corresponds to 300/200mg TDF/FTC, 50mg DTG and 400mg EFV. The colored lines depict the median predicted PEP efficacy, whereas the dark- and light grey areas present the inter-quartile range and the 95% confidence range, respectively. Dashed horizontal lines indicate 90% prophylactic efficacy.

### Third drug may be added later, if TDF/FTC is initiated quickly

In many settings, all three drugs may not be available within reasonable time. However, TDF/FTC may be readily available to individuals who already used, or have access to PrEP. We investigated whether prompt PEP initiation with TDF/FTC and later addition of a third drug may effectively prevent infection (schematic in Fig 2A). Reading Fig 2B-E bottom-to-top, indicates that adding DTG or EFV to a TDF/FTC backbone increases PEP efficacy (lowest row: TDF/FTC only) and that earlier addition of the third drug results in greater efficacy (top row). Reading Fig 2B-E horizontally (left-to-right), indicates that the earlier TDF/FTC is initiated, the better. For the three drug combinations, a ‘window of opportunity’ arises, where the PEP efficacy exceeds 95%. For TDF/FTC + DTG the duration of PEP strongly impacts on its prophylactic efficacy (compare panels B and D in Fig 2), whereas the impact is less strong for PEP with TDF/FTC + EFV, which is already efficient for 2 weeks PEP. The simulations highlight that if TDF/FTC is available within 12-24 hours, the third drug should be added in less than a weeks time and PEP should preferably be taken for 28days from the first TDF/TFC dose.

**Fig 2.**
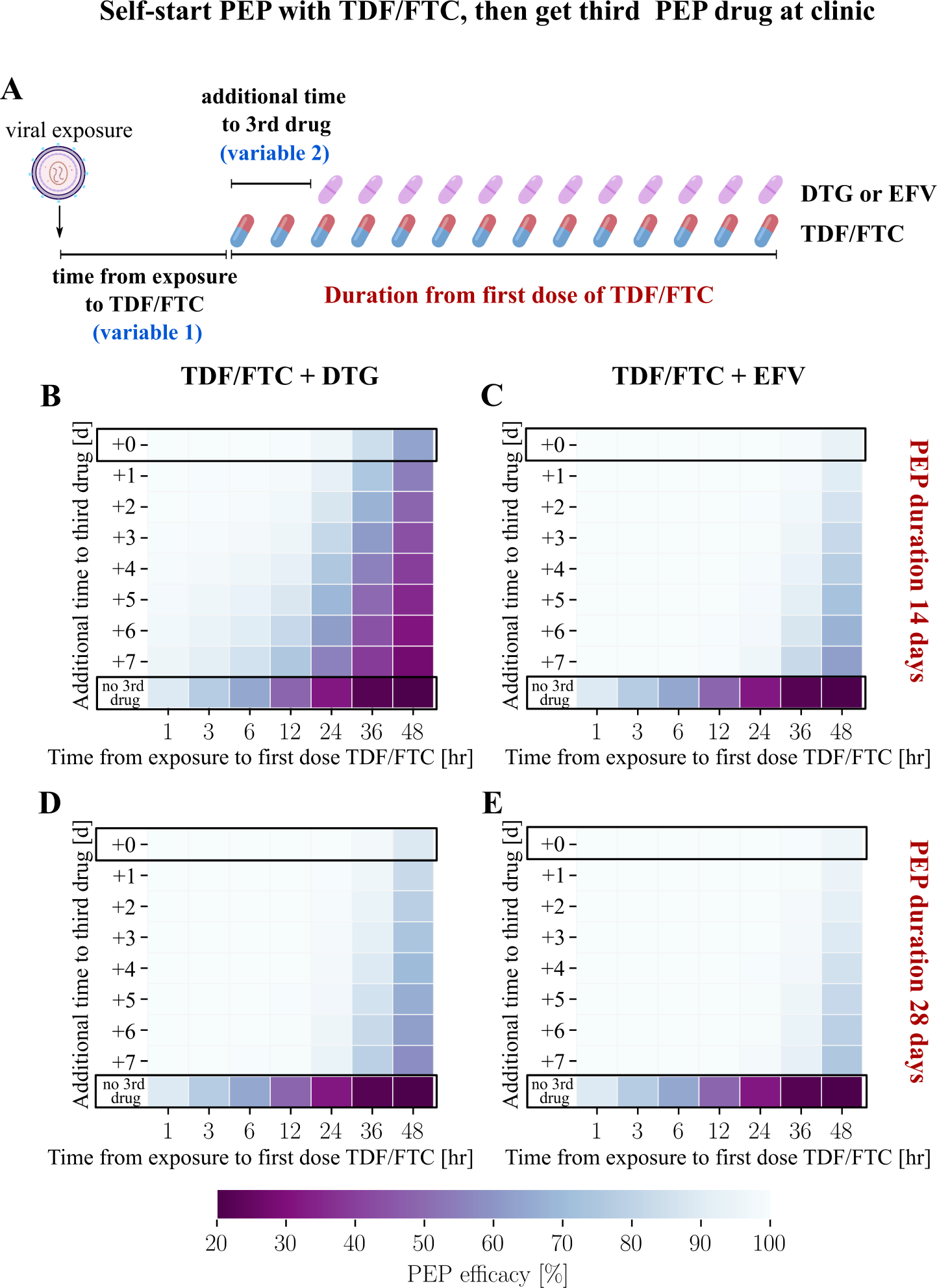
Efficacy of TDF/FTC-based PEP with delayed initiation of TDF/FTC and further delay of the third drug. A:Schematic of the dosing regimen. For the drug combinations TDF/FTC + DTG and TDF/FTC + EFV, PEP efficacy was computed for virus exposures occurring within 1 to 48 hours before the first dose of TDF/FTC. The third drug was then added to the PEP regimen 1 to 7 days after the the first dose of TDF/FTC. B: PEP efficacy for the drug combination TDF/FTC + DTG, PEP duration was 14 days from the first dose of TDF/FTC. C: Corresponding PEP efficacy for TDF/FTC + EFV. D: PEP efficacy for TDF/FTC + DTG when taken for 28 days after the first TDF/FTC dose. E: Corresponding PEP efficacy for TDF/FTC + EFV. In panel B-E, the top row outlined in black denotes the scenario where the third drug is immediately added to the TDF/FTC backbone; the bottom row represents the scenario where no third drug was added to the TFC/TDF backbone.

### Previous PrEP can boost subsequent PEP efficacy and widen the ‘window of opportunity’

The pharmacologically active components of TDF and FTC (TFV-DP and FTC-TP respectively) are built-up slowly within HIV target cells [13,14,24,30], which limits the ‘window of opportunity’ for 2-drug (TDF/FTC) PEP (Fig 1C) and necessitates almost instantaneous PEP initiation to achieve sufficient efficacy. However, TFV-DP and FTC-TP may persist for many hours even after a single, or a few dosing events. To assess the combined impact of earlier TDF/FTC PrEP intake with PEP, we investigated the efficacy of PEP following an ‘on-demand’ (2-1-1) PrEP regimen [31] (schematic in Fig 3A). In our simulations, viral exposure occurs 2 (panels B, F), 3 (panels C, G) or 7 (panels D, H) days after the last PrEP ‘on demand’ dose. A two- or three drug PEP regimen is then initiated within 0–72 hours post virus exposure (x-axis) and continued for either for 7 (panels B-E) or 28 days (panels F-I).

**Fig 3.**
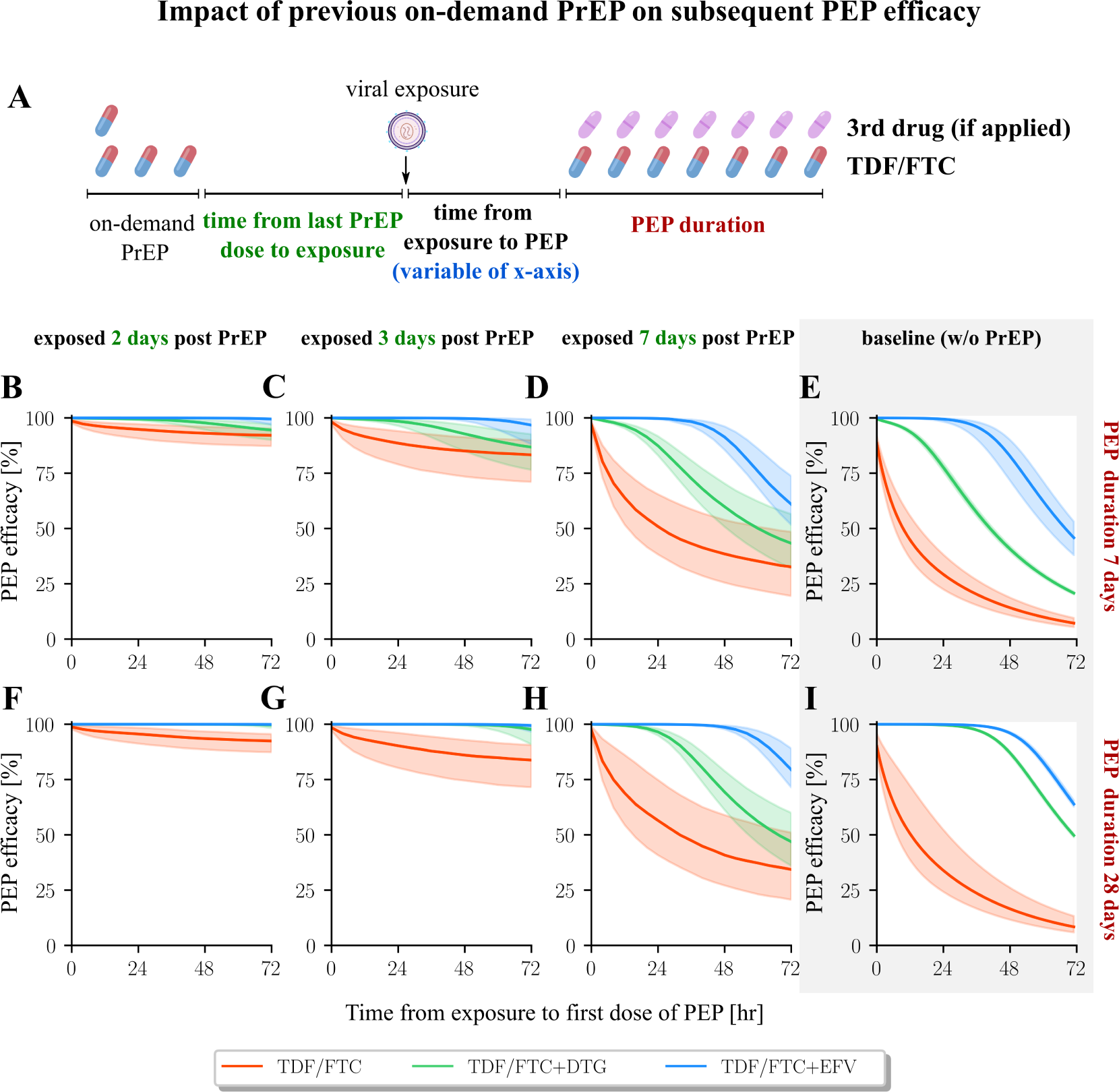
PEP Efficacy Following On-Demand PrEP. A: Schematic of the dosing regimen. Truvada was initially administered as “on-demand” PrEP (2-1-1), followed by viral exposure after a certain period. Subsequently, the PEP regimen was initiated after various time intervals, potentially incorporating a third drug. B-D: the efficacy profiles for PEP with overall duration of 7 days, and the exposure occurred 2 days, 3 days and 7 days after the on-demand PrEP, respectively. F-H: the efficacy profiles for PEP with overall duration of 28 days. E&I: PEP efficacy of baseline scenario without preceeding PrEP. All computations were performed on 1000 virtual patients. The daily dose for each drug corresponds to 200 mg FTC, 300 mg TDF, 50 mg DTG, and 400 mg EFV. The colored lines represent the median efficacy value in cases where PEP was initiated at the respective time point along the x-axis. The shaded areas depict the quantile range of prophylactic efficacy.

Reading Fig 3B–D and Fig 3F–H left-to-right shows that if the last PrEP-on-demand dosing event was 7days ago, the added benefit of earlier PrEP-on-demand on subsequent PEP efficacy had almost vanished, compare to Fig 3E and Fig 3I (no preceding PrEP). However, if PrEP- on-demand was taken less than 7days prior to virus exposure, it increases subsequent PEP efficacy, as residual FTC-TP and TFV-DP concentrations may be present that either prevent infection in some individuals, delay sero-conversion [32, 33], or result in a ‘pre-loading’ of drug concentrations for subsequent PEP. For example, if on-demand-PrEP was stopped 2days prior to virus exposure, subsequent PEP with TDF/FTC may be *>*90% efficient, even when initiated within three days, Fig 3B,F. For the three-drug PEP regimen we observed a *>*90% efficacy when initiated within three days after viral exposure and taken for *>*7days, Fig 3C,G. Overall, we observe that earlier PrEP combined with subsequent PEP can increase efficacy.

Next, we investigated the concomitant impact of preceding daily PrEP with 1–7 average doses per week, stopped 2days before viral exposure, in conjunction with subsequent 2-drug or 3-drug PEP, initiated 2-, 3-, or 7days after virus exposure and taken for 28days (schematic in Fig 4A). As controls, we performed simulations without earlier PrEP (grey-shaded areas), as well as PrEP-only simulations (empty box plots) in Fig. 4B-D. Our simulations confirm the combined action of PrEP and PEP: Earlier PrEP boosts the efficacy of PEP, if PEP is initiated 2-, or 3days post-exposure, Fig. 4B–C: Compared both to ‘no-PrEP’ (grey-shaded areas), as well as ‘no-PEP’ (empty boxplots), prophylactic efficacy is increased for the PrEP+PEP combination. However, PEP does not offer any additional protection when initiated 7days post-exposure (compare empty- vs. coloured box-plots in Fig 4D). Interestingly, our model predicts that PrEP-only with 100% adherence offers *>* 90% protection, when stopped 2 days before virus exposure (empty bars in Fig. 4D). Also, for the PrEP+PEP combination we observe *>* 95% protection, if 4/7 doses of earlier PrEP were taken and 3-drug PEP was initiated 3days post exposure. For comparison, PEP-only offers only 50% (TDF/FTC/DTG) and 65% (TDF/FTC/EFV) protection if initiated 3days post exposure (Fig. 1C and Fig. 4C). If PEP is initiated 2days post-exposure, preceding PrEP may lift prophylactic efficacy from 90% (TDF/FTC/DTG) and 95% (TDF/FTC/EFV) to almost complete protection, if adherence during preceding PrEP was 2/7 doses (EFV) vs. 3/7 (DTG).

**Fig 4.**
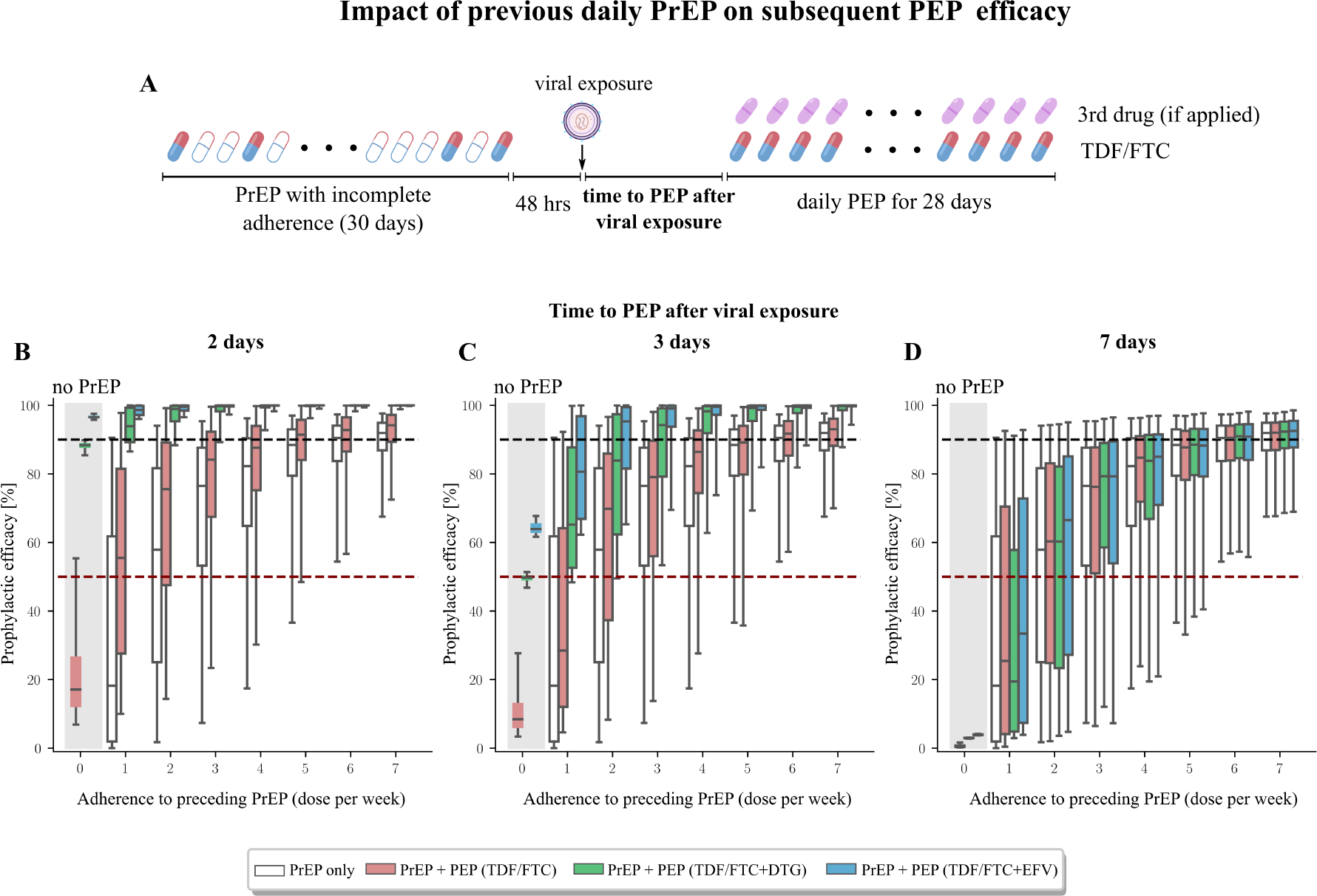
Predicted efficacy of once-daily PEP, in cases where PrEP was recently taken. A: Schematic of dosing regimen: PrEP with incomplete, variable levels of adherence was taken and stopped 24hours before virus exposure. PEP with either TDF/FTC, or TDF/FTC + DTG or EFV was then initiated after a variable delay and taken for 28 days. PEP efficacy is calculated with regards to preceding PrEP adherence, as well as delay in PEP initiation. B-D: Computed prophylactic efficacy for the distinct PrEP+PEP regimen, if PEP was initiated 2, 3, or 7 days post-exposure and taken daily for 28days. The grey-shaded area indicates PEP efficacy, with no prior PrEP, while empty boxplots highlight the prophylactic effect of preceding PrEP, without subsequent PEP. Boxplots show the median, interquartile ranges and whiskers encompass the 95% confidence interval.

Lastly, we tested scenarios in which the probability of PEP adherence declined substantially over time. We modelled PrEP with incomplete adherence 48 hours prior to virus exposure (schematic: Fig 5A). We further assumed a substantial decrease in PEP-adherence after 7days, Fig 5B. Overall, compared to a full 28days PEP regimen simulated in Fig 4B, we can see a drug-specific decline in efficacy that is clearly seen in simulations without preceding PrEP (grey shaded area in Fig 4C): Two-drug TDF/FTC is already quite inefficient (*<* 20%) when initiated 2, 3 or 7days post exposure and hence poor PEP-adherence marginally impacts (grey-shaded areas in Fig 4B–D vs Fig 5C–E). In contrast, for the three-drug combinations, we see that poor PEP-adherence negatively impacts on prophylactic efficacy (compare shaded areas in Fig. 4B–C with Fig. 5C–D). However, if ≥4/7 doses of earlier PrEP were taken and subsequent 3-drug PEP was initiated ≤ 3days post-exposure, we predicted that prophylactic efficacy may exceed 95%. In summary, we observe that preceding PrEP can substantially boost subsequent PEP efficacy for all drug regimen, and ‘buy time’ with regards to PEP initiation, particularly if preceding PrEP adherence was good (≥ 3 doses/week).

**Fig 5.**
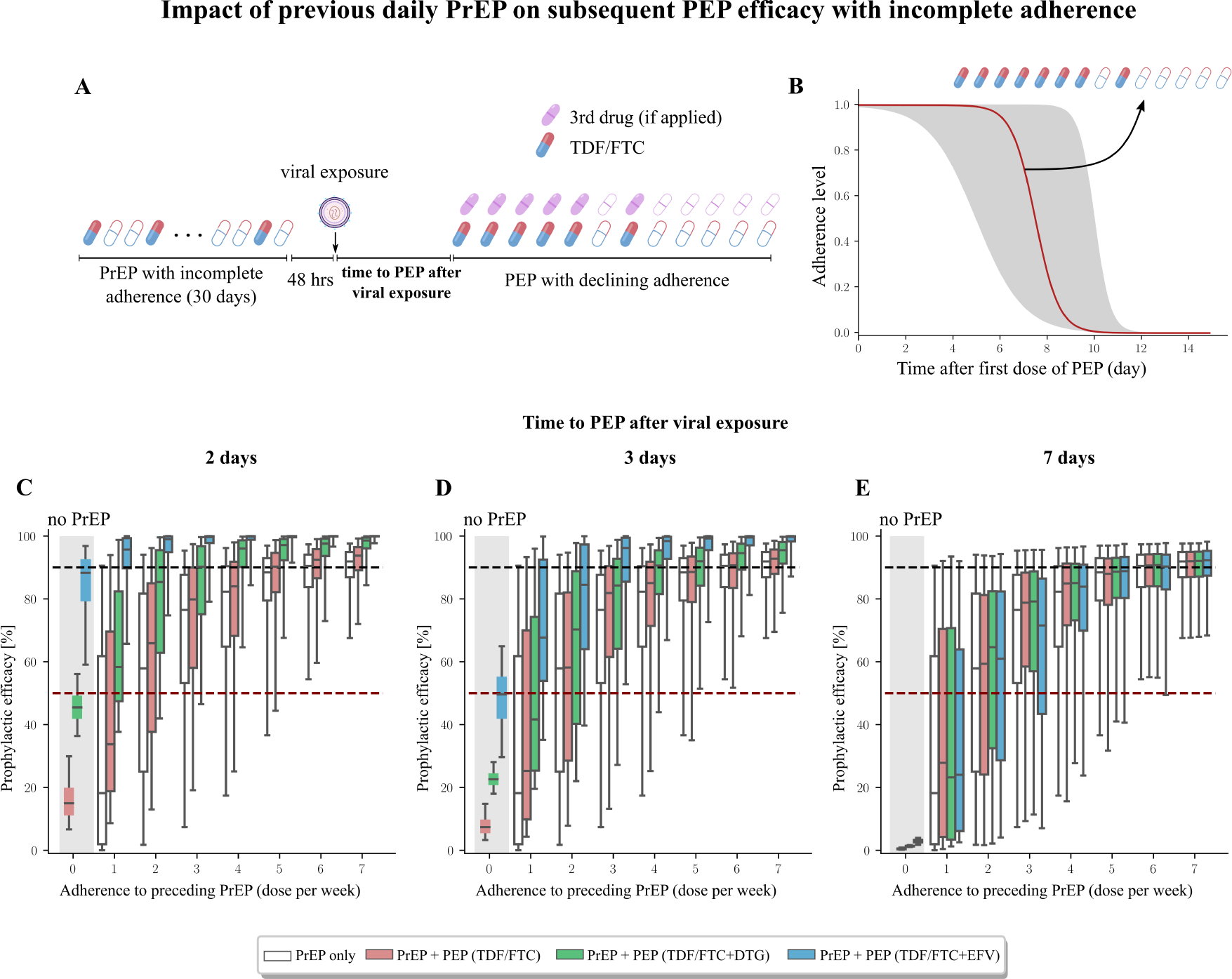
Predicted efficacy of PEP with strongly declining adherence, in cases where PrEP was recently taken. A: Schematic of dosing regimen: PrEP with incomplete, variable levels of adherence was taken and stopped 24 hours before virus exposure. PEP with either TDF/FTC, or TDF/FTC + DTG or EFV was then initiated after a variable delay and adherence strongly decreases over time. PEP efficacy is calculated with regards to preceding PrEP adherence, as well as delay in PEP initiation. B: Simulated PEP adherence probability with a half maximum at 7 days post PEP initiation. C–E: Computed prophylactic efficacy for the distinct PrEP+PEP regimen, if PEP was initiated 2, 3, or 7 days post-exposure and adherence declined substantially after 7 days. The grey-shaded area indicates PEP efficacy without prior PrEP, while empty boxplots highlight the prophylactic effect of preceding PrEP, without subsequent PEP. Boxplots show the median, interquartile ranges and whiskers encompass the 95% confidence interval.

## Discussion

We evaluated the impact of delays in ‘time to PEP’, PEP duration and PrEP-to-PEP transition, based on a combined model of drug-specific pharmacokinetics and viral dynamics. Our modelling by-and-large confirms recent UK, US and WHO guidelines on PEP [6–8], which recommend to combine a TDF/FTC backbone with a third drug, initiate PEP as early as possible and to take it for 28days. Moreover, our simulations indicate that early PEP initiation after suspected virus exposure denotes the most critical parameter. For TDF/FTC two-drug PEP, instantaneous initiation would be required, which may be infeasible. Adding a third drug to the TDF/FTC-backbone ‘buys time’. However, protection may still be incomplete (Fig 1C), if a three drug PEP was initiated 72hours post virus exposure and taken for 28days. The duration of PEP was a less sensitive parameter for EFV, compared to DTG. The latter is contributed to the half-life (*t*_1*/*2_) of the drugs, with DTG having a relatively short half-life (*t*_1*/*2_ = 13.5 − 15.9h [15] compared to EFV (*t*_1*/*2_ = 40 − 55h) [27]. The long half-life may increase the likelihood that virus is cleared before the drug is washed out of the body. While early PEP initiation may be particularly difficult in settings with less established health infrastructure, we simulated scenarios, in which PEP may be initiated with available TDF/FTC and later intensified with a third drug. Overall, if TDF/FTC can be initiated within less than 24hours, the third drug may be added, as soon as it becomes available (Fig 2), on condition that PEP is taken 28days. We found that preceding TDF/FTC-based PrEP can substantially boost PEP efficacy, if stopped no more than 3 days before suspected virus exposure (Fig. 3), or taken at 4/7 days on average (Fig 4–5). Thus, individuals taking PrEP up to the time of exposure (−3days) could re-initiate the regimen and may add a third drug when it becomes available. The combined effects of PrEP+PEP in this scenario indicate synergy, which could arise from the fact that previous PrEP delays initial viral replication [33], or pre-loads drug levels for subsequent PEP. Our simulations further highlighted that daily PrEP-only with 100% adherence may provide *>* 90% protection, if stopped no more than 48hours before exposure (Fig. 4D and Fig. 5E). Essentially, this observation is backed by the long pharmacokinetic halflifes of TFV-DP and FTC-TP in PBMCs, in the range of 4–7 and 1-2.2 days respectively [34–38].

Our work has a number of limitations: Foremost, there is a lack of data that could be inputted into the model, due to a lack of clinical research into PEP. To strengthen the model further clinical trials with clinically relevant endpoints may be required.

Our simulations refer to exposure with ‘wild type’ viruses, whereas NNRTI drug resistance, which may amount to 10 − 20% of transmitted viruses in Africa and the Americas [39, 40] may severely diminish EFV-based PEP efficacy [27] and thus the suitability of EFV as a PEP component. Notably, while we include EFV in our analysis to explore the impact of a 3rd drug components with very high molecular potency [41], we are not advocating EFV for PEP as it is contraindicated both for psychological side effects and low risk of serious liver toxicity. However, while some clinical trials suggest superiority of integrase inhibitors (DTG over EFV) [42–45] with regards to ‘time to viral load suppression’, we would like to emphasize that viral load kinetics decay more strongly for integrase inhibitors, merely because they inhibit a later stage of the viral replication cycle and not because of superior efficacy (or potency) [46–48]. Hence, the current preference for integrase inhibitors in PEP regimen should be motivated by tolerability and low prevalence of drug resistance rather than alleged efficacy. We did not investigate ritonavir-boosted protease inhibitors lopinavir (LPV/r) or atazanavir (ATV/r) as third drug components in our model [7]. While these compounds have high molecular potency [41] we expect PEP efficacy to be similar to EFV. However, previous work suggests very steep dose-response curves for LPV/r and ATV/r, implying that prophylactic effect may rapidly drop in case of incomplete PEP-adherence, or discontinuation [49]. In our model, we assume that the effect of the considered drugs is associated with systemic drug levels. Both EFV and DTG are lipophilic drugs that can rapidly cross cellular membranes by passive diffusion, such that their unbound drug concentration in plasma strongly correlates with effect-site concentrations (‘free drug hypothesis’ [50, 51]). With regards to TDF/FTC, their phosphate moieties (TFV-DP/FTC-TP) in peripheral blood mononuclear cells (PBMCs) were used as an effect marker, since our recent work [12] indicated strong correlation with effect, whereas concentrations in tissue homogenates were not predictive regarding prophylactic efficacy.

With regards to pharmacodynamics, we simulated synergistic effects between TFV-DP and FTC-TP, based on recent results [28] and assumed that the direct antiviral effects of DTG and EFV are additive to the TDF/FTC-backbone.

In our simulations, we modelled viral challenges after sexual exposure (receptive vaginal intercourse). Notably, the majority of non-occupational PEP is administered after potential sexual exposure (PEPSE) [52] and women denote the major HIV risk group [53]. Occupational virus exposures, through e.g. needle-stick injuries during healthcare procedures may lead to the translocation of larger amounts of viruseswhich may negatively impact on prophylactic efficacy [30]. Thus, our predictions may be optimistic regarding occupational exposures with patient blood.

## Conclusions

Our modelling suggests that ‘time to PEP’ denotes the most critical parameter. Three-drug PEP, preferably initiated no later than 48hours after virus exposure, and taken for 28days remains the optimal regimen. Three-drug PEP for 14days is less efficient than 28days and 2-drug (TDF/FTC) PEP only has high efficacy, if started within one hour after exposure. Self-start 2-drug (TDF/FTC) PEP with a subsequent addition of a 3rd drug in the clinic works better than not self-starting. Lastly, previous PrEP intake *<* 7days prior to virus exposure boosts subsequent PEP efficacy and may widen the window period for ‘time to PEP’ past 72hours.

## Data Availability

All data produced are available online at https://github.com/KleistLab/PEP

## Authors’ contributions

L.Z. and M.v.K. wrote the manuscript with help from J.F. and S.C. L.Z. and M.v.K. designed the research. L.Z. performed the research and L.Z., M.v.K., J.F. and S.C. analyzed the data.

## Acknowledgements Funding

M.v.K. acknowledges funding from the German ministry for science and education (BMBF), grant number 01KI2016, from the DFG research center MATH+, as well as “Sonderforschungsmittel” (SoFo) provided through the Robert-Koch Institute. The funders had no role in the design of the study or the decision to publish.

## Data Availability Statement

All data and computational codes are available at https://github.com/KleistLab/PEP

## Supporting Information

### Supplementary Text S1: Viral dynamics of HIV

We employ a viral dynamic model of HIV [17, 18], which contains six viral compartments: free infectious viruses V, early infected cells, i.e. T-cells T_1_ and macrophages M_1_, productively infected cells T_2_ and M_2_, and the latently infected T-cells T_L_. As depicted in Fig S1, the dynamics can be defined by 15 reactions whose reaction propensities are denoted as *a*_1_ through *a*_15_. The drug classes that are investigated in this work are also integrated in this viral dynamics. Equations (S1)–(S15) define the concrete propensity of reaction.

**Figure S1.**
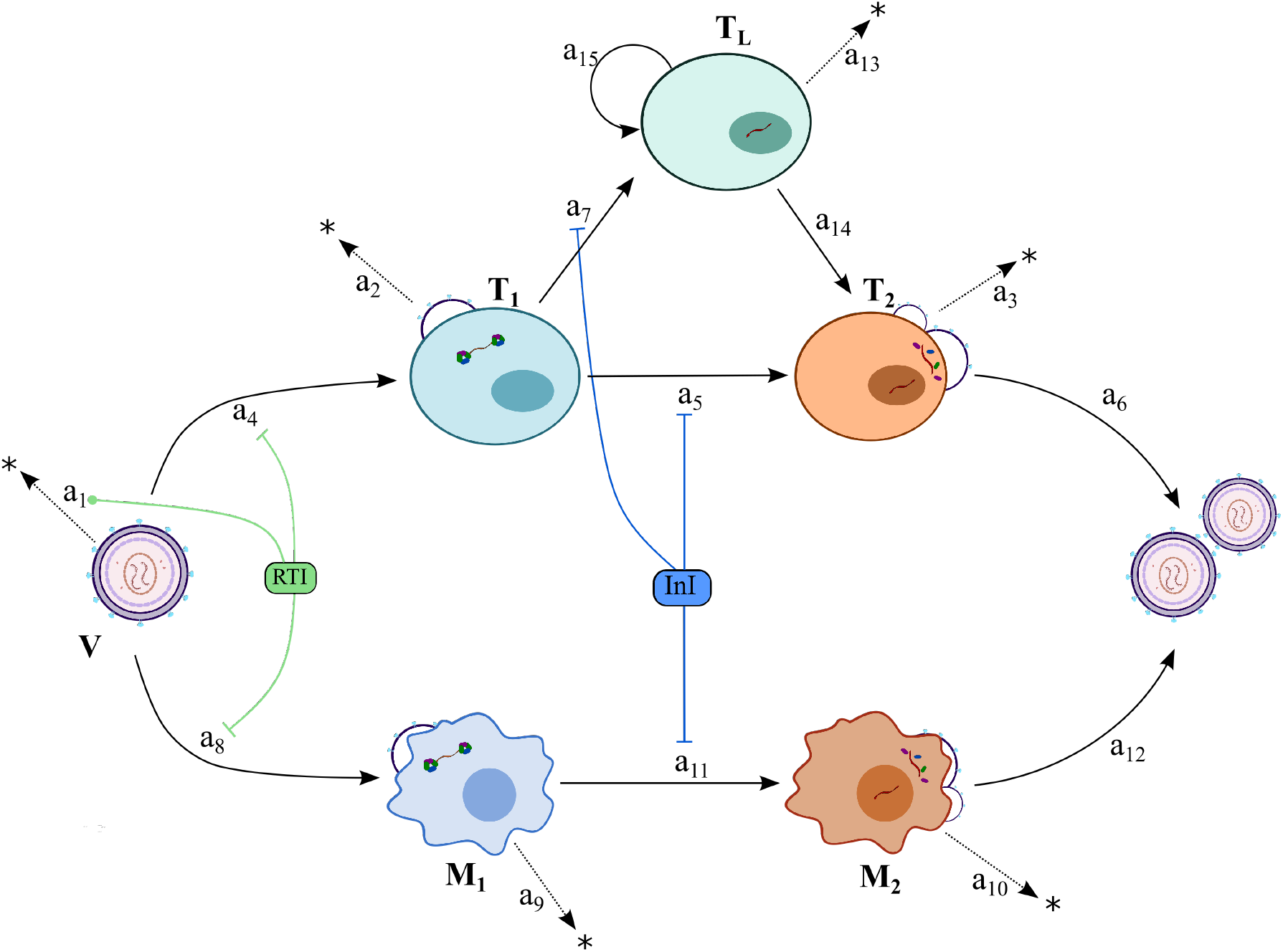
Illustration of the viral dynamic model and the interference mechanisms of different drug classes. Free infectious viruses V can infect target cells and create early infected T-cells T_1_ and macrophages M_1_ after successful infection. In early infected cells, the viral DNA can become integrated into the host genome, creating late infected cells T_2_ and M_2_, which are able to release new viruses. Early infected T-cells T_1_ can become latently infected, i.e. the cells will transition into a resting state, denoted as T_L_. The latent infected T-cells can replicate itself or be reactivated and turn into T_2_ cells. Viral compartments can also be eliminated. *a*_*j*∈1…15_ denotes the propensity of each reaction (see equations (S1)-(S15)). RTI: reverse transcriptase inhibitor; InI: integrase inhibitor.

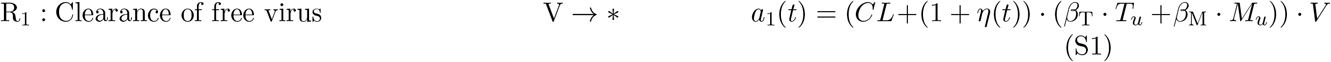

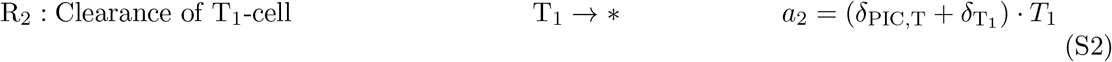

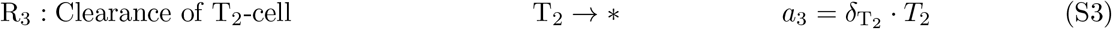

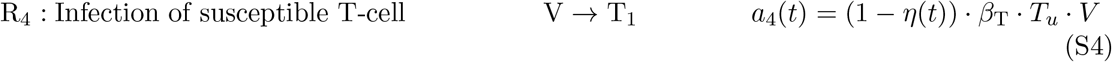

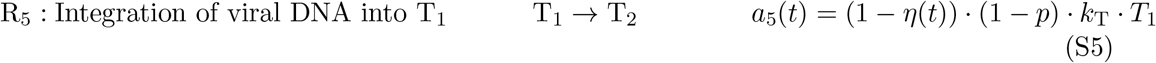

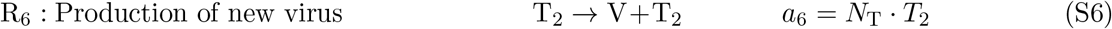

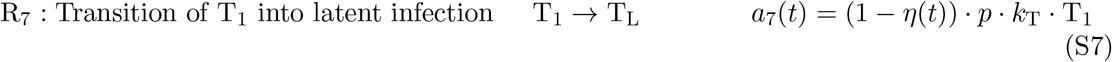

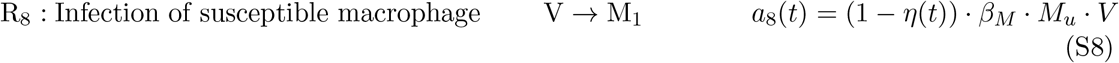

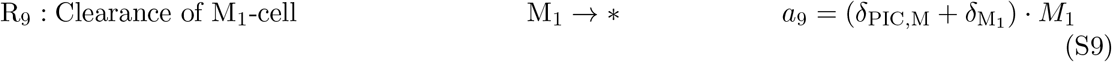

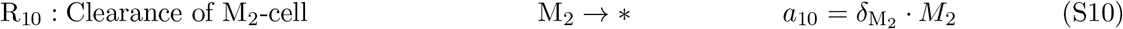

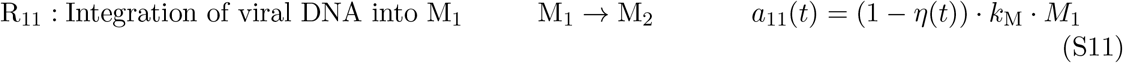

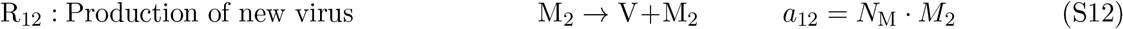

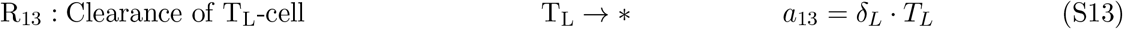

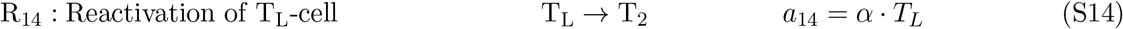

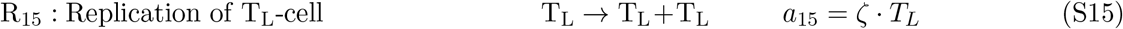

Here we assume a continuous virus production model, where T_2_ and M_2_ cells will produce viruses continuously until they are eliminated. The used parameters except for the replication rate of T_L_ are listed in [18], Table 1 therein. The T_L_ replication rate *ζ* is adopted from [54].

### Supplementary Text S2: numerical approach for PEP efficacy

In the *Methods* section we defined the prophylactic efficacy of a given prophylactic regimen, as the reduction in infection probability *per exposure*. Since extinction probability is the complement of infection probability, we can compute the extinction probability by adopting a recently developed numerical approach [19]. Based on the viral dynamics model in Fig S1, the extinction probability *P*_*E*_ of each single viral compartment can be derived as follows:

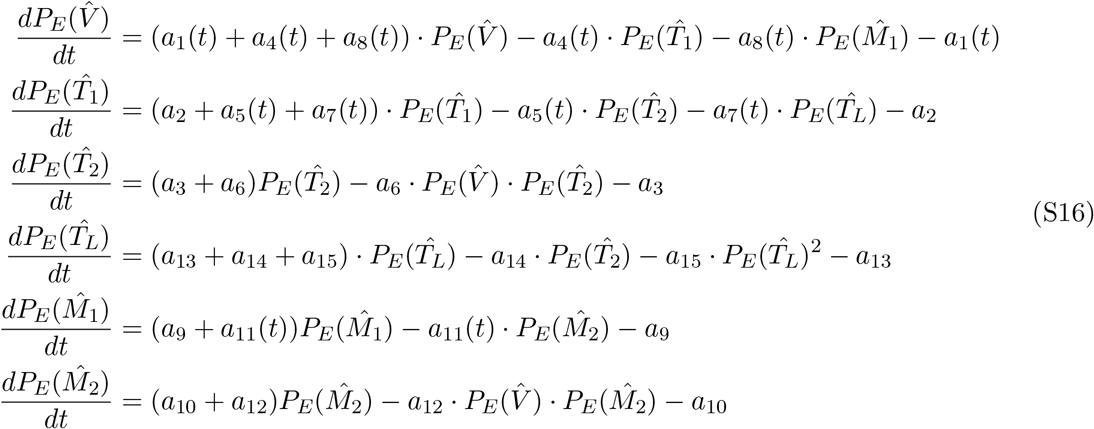

The time-dependent reaction rates are given in eqs (S1)–(S15). The system of ordinary differential equations (S16) is solved backwards using standard ODE solvers, as outlined in [19].

